# Preparedness and response to Pediatric CoVID-19 in European Emergency Departments: a survey of the REPEM and PERUKI networks

**DOI:** 10.1101/2020.04.28.20075481

**Authors:** Silvia Bressan, Danilo Buonsenso, Ruth Farrugia, Niccolo’ Parri, Rianne Oostenbrink, Luigi Titomanlio, Damian Roland, Ruud G. Nijman, Ian Maconochie, Liviana Da Dalt, Santiago Mintegi

**Author notes:** Correspondence to: Silvia Bressan MD, PhD, Department of Woman’s and Child’s Health – Division of Pediatric Emergency Medicine, University of Padova, Italy; /3570, (preferred). **Meetings:** This work has not been presented to any meeting. **Grant funding:** No external source of funding supported the present work. **Conflicts of interest:** All the authors have no conflicts of interest to disclose.

## Abstract

**Study objective:** We aimed to describe the preparedness and response to the COVID-19 pandemic in referral EDs caring for children across Europe.

**Methods:** We did a cross-sectional point prevalence survey, which was developed and disseminated through the pediatric emergency medicine research networks for Europe (REPEM) and the United Kingdom and Ireland (PERUKI). We included a pre-determined number of centers based on each country population: five to ten EDs for countries with > 20 million inhabitants and one to five EDs for the other countries. ED directors or named delegates completed the survey between March 20th and 21st to report practice in use one month after the outbreak in Northern Italy. We used descriptive statistics to analyse data.

**Results:** Overall 102 centers from 18 countries completed the survey: 34% did not have an ED contingency plan for pandemics and 36% had never had simulations for such events. Wide variation on PPE items was shown for recommended PPE use at pre-triage and for patient assessment, with 62% of centers experiencing shortage in one or more PPE items. COVID-19 positive ED staff was reported in 25% of centers. Only 17% of EDs had negative pressure isolation rooms.

**Conclusion:** We identified variability and gaps in preparedness and response to the COVID-19 epidemic across European referral EDs for children. Early availability of a documented contingency plan, provision of simulation training, appropriate use of PPE, and appropriate isolation facilities emerged as key factors that should be optimized to improve preparedness and inform responses to future pandemics.

## INTRODUCTION

### Background

Ever since the first human cases of the novel coronavirus were reported in Wuhan, Hubei Province in China in December 2019, the Coronavirus Disease 2019 (COVID-19) pandemic has spread rapidly across the world.^1^ The epidemic in Europe initially centerd around Northern Italy where there was a steep rise in the number of cases and case fatalities from February 20^th^ onwards.^2^ While European countries were deciding upon or were enacting containment measures of varying degrees, the infection continued to spread across the continent with devastating impact on health systems, the economy and the society at large.

It is now more crucial than ever before that the emergency department (ED), as the entry point to hospital care, is prepared to manage high risk patients in an efficient and safe way, from triage to final disposition. The ED should respond to the epidemic surge in agreement with hospital contingency plans and guidelines from local and national health authorities,^3^ also drawing on the experience of other countries.^4^

With the pandemic unfolding throughout Europe, pediatric emergency physicians liaised through their European Society - The European Society of Emergency Medicine (EuSEM) Pediatric Emergency Medicine (PEM) Section, and European Pediatric Emergency Medicine research networks - Research in European Padiatric Emergency Medicine (REPEM) and Pediatric Emergency Research United Kingdom and Ireland (PERUKI), to share experiences and resources.^11–13^ This dialogue highlighted the differences and challenges of ED preparedness, surge capacity and management of pediatric suspected/confirmed COVID-19 cases between countries. Although European countries greatly differ in their culture, legislation, health care systems, and territorial organization, physicians working in the frontline of pediatric emergencies strongly advocate for generalizable guidance to enhance preparedness and readiness to pandemic emergencies across the whole age spectrum, to better face COVID-19 and possible future pandemics.

### Importance

Even though it has now become apparent that children are affected less frequently and with a much more benign disease spectrum than adults,^5,6^ appropriate management of suspected and confirmed cases and their families are essential throughout all levels of health care systems.^7,8^ EDs should also maintain the quality of care provided to children presenting with serious illnesses or accidents not related to the pandemic. Pathway and protocols need to be in place to ensure that rapid appropriate care is provided to suspected COVID-19 children, while avoiding delay in care of non-COVID-19 patients.^9^ In addition, it will be paramount to ensure patients and staff are protected from the infection and with as little exposure as possible.^10^ It has also become recognised that children may present with conditions not linked to COVID-19 but some, when admitted for that condition, are found to have COVID-19 positive swabs as an incidental finding. This may be a feature with the more widespread dissemination of COVID-19 throughout the population.

### Goals of this investigation

Hence, we developed a structured point prevalence survey to describe the preparedness and response to the COVID-19 pandemic, including strengths and challenges, in European referral EDs for children within the REPEM and PERUKI networks. The secondary objective was to summarize the lessons learnt, which can be generalizable across countries.

## METHODS

### Survey design and setting

This was a cross-sectional point prevalence study to describe the preparedness and response to the COVID-19 pandemic and to explore common themes in lessons learnt from the pediatric emergency field across Europe. The survey was developed in English by the lead author and then underwent several rounds of review by the research team. The survey was distributed through the REPEM network,^12^ a research collaborative consisting of Pediatric EDs (PEDs) and EDs of general hospitals with a separate pediatric section, serving as referral centers for children and also the sites affiliated to the executive committee members of PERUKI. For each European Country a country lead was identified to disseminate the survey to centers meeting the above criteria. We also included Isrel as a European associated country, as Isrel has been part of the REPEM network since its foundation.^14^ To ensure balanced representativeness of participating countries and feasibility of the study, the research team agreed to include a pre-determined number of centers based on the population of participating countries. For countries with more than 20 million inhabitants the country leads had to include five to ten EDs; for countries with less than 20 million inhabitants this was one to five EDs. We defined ED directors or named delegates as most suitable persons to complete the survey, and they were asked to complete one survey for each participating center. The survey was open on March 20^th^ and 21^st^ with specific instructions to respond reporting information available and practice in use on March 20^th^ 2020. Survey responses were collected in REDCap, a validated online data collection system.^15^ Respondents were asked to state their country of residency, but it was not mandatory to give the name of their hospital. Each country-lead recorded the name of the invited and participating hospitals. Country leads communicated to the principal investigator the number of centers that completed the survey, without disclosing the hospitals’ identities, ensuring the number of completed surveys per country matched the number of centers that actually completed the online survey.

### Survey content

A first survey was completed by country-leads to reflect the national situation of the COVID-19 pandemic as of March 20 2020. The country lead survey included questions on range of COVID-19 cases (total and pediatric) per country; the date of identification of the first COVID-19 cases in the country; and the type of containment measures enforced in their country. Data on range of confirmed cases per country, as well as deaths, were cross checked with the European center for Disease Control and worldometers websites on March 21^st^, to ensure complete update of data up to March 20^th^.^16,17^ The formal study survey completed by each participating center focused on organizational and operational aspects of preparedness and response including contingency plan, training, screening criteria for suspected cases, capacity, personal protective equipment, ED infection control measures and management of patients, health professionals safety and sustainability of care, resources found useful to prepare the ED for management of pediatric cases. All answers had to be provided as per practice on March 20^th^.

### Statistical analysis

Descriptive statistics were used to analyse the data. Association between categorical variables was tested by means of chi square or Fisher exact test as appropriate.

Data were analysed using Stata (version 13, StataCorp, College Station, Texas, USA). P-values were considered significant if P was less than 0·05.

### Ethics

This survey accessed clinicians via a research collaborative to assess their departmental practice and therefore did not require formal ethics review, as per consultation with the ethics committee of the University Hospital of Padova, Italy. Consent was implied by participation.

## RESULTS

A total of 102 centers from 18 countries completed the survey. The survey was completed by the ED director in 48% of cases and by their delegate in 52%. The number and characteristics of participating centers and the range of COVID-19 confirmed cases per country as of March 20^th^ is reported in **Table 1**. The majority of participating EDs were tertiary-care PEDs (75%) and most centers have a pediatric yearly census > 10,000 visits per year (89%). Only few confirmed COVID-19 pediatric patients, if any, were seen in participating EDs. Containment measures enforced in participating countries as of March 20^th^ were highly variable and their number was not associated with the infection spread (**Table 1S; Figure 1S – Supplementary material**). Measures less often taken were the most restrictive, namely the closure of non-essential commercial activities (67%); the closure of land borders (50%) and the prohibition of any travel not-related to health or food shopping needs (50%). A summary of criteria for suspected COVID-19 case in use at each participating ED is provided in **Table 2S-Supplementary material**. Definition criteria had changed over time in 90% of centers, reflecting the very dynamic adjustments made to face a rapidly evolving crisis. At the time of survey, any child with flu-like illness or fever was considered as a suspected COVID-19 infection in 67% of centers. Countries with a longer time interval from their first COVID case were significantly more likely to adopt this definition.

**Table 1.** Characteristics of participating countries and centers

|  | Belgium | Denmark | Estonia | France | Germany | Iceland | Ireland | Israel | Italy | Latvia | Lithuania | Malta | Netherlands | Portugal | Spain | Sweden | Switzerland | UK |
| --- | --- | --- | --- | --- | --- | --- | --- | --- | --- | --- | --- | --- | --- | --- | --- | --- | --- | --- |
| Number of centers | 7 | 5 | 3 | 15 | 13 | 1 | 4 | 6 | 11 | 1 | 2 | 2 | 2 | 5 | 9 | 3 | 6 | 7 |
| ED Setting |  |  |  |  |  |  |  |  |  |  |  |  |  |  |  |  |  |  |
| - Tertiary care PED of standalone hospital | 0 | 0 | 1 | 6 | 4 | 1 | 2 | 1 | 5 | 1 | 0 | 0 | 0 | 2 | 3 | 1 | 3 | 1 |
| - Tertiary care PED in a hospital for adults and children | 2 | 3 | 1 | 7 | 8 | 0 | 0 | 5 | 6 | 0 | 1 | 0 | 1 | 1 | 5 | 1 | 1 | 3 |
| - Referral general ED with pediatric section * | 5 | 2 | 1 | 2 | 1 | 0 | 2 | 0 | 0 | 0 | 1 | 1 | 1 | 0 | 0 | 0 | 2 | 3 |
| - Other # | 0 | 0 | 0 | 0 | 0 | 0 | 0 | 0 | 0 | 0 | 0 | 1 | 0 | 2 | 1 | 1 | 0 | 0 |
| Pediatric age limit |  |  |  |  |  |  |  |  |  |  |  |  |  |  |  |  |  |  |
| - up to 14 years of age | 0 | 0 | 0 | 0 | 0 | 0 | 0 | 0 | 4 | 0 | 0 | 0 | 0 | 0 | 4 | 0 | 0 | 0 |
| - up to 15 years of age | 2 | 0 | 0 | 3 | 0 | 0 | 1 | 0 | 2 | 0 | 0 | 0 | 0 | 0 | 2 | 0 | 0 | 1 |
| - up to 16 years of age | 3 | 0 | 0 | 3 | 0 | 0 | 3 | 0 | 1 | 0 | 0 | 1 | 0 | 0 | 1 | 0 | 5 | 6 |
| - up to 18 years of age | 2 | 5 | 3 | 9 | 13 | 1 | 0 | 6 | 4 | 1 | 2 | 1 | 2 | 5 | 2 | 3 | 1 | 0 |
| ED pediatric yearly census (visits/year) |  |  |  |  |  |  |  |  |  |  |  |  |  |  |  |  |  |  |
| - < 10,000 | 1 | 2 | 2 | 0 | 2 | 0 | 0 | 0 | 0 | 0 | 0 | 1 | 2 | 0 | 0 | 1 | 0 | 0 |
| - 10,000 -15,000 | 4 | 0 | 0 | 0 | 0 | 0 | 0 | 0 | 1 | 0 | 1 | 0 | 0 | 0 | 0 | 1 | 0 | 1 |
| - 15,000 -25,000 | 2 | 3 | 0 | 0 | 9 | 1 | 1 | 1 | 4 | 0 | 0 | 1 | 0 | 1 | 2 | 1 | 2 | 0 |
| - 25,000 -50,000 | 0 | 0 | 0 | 11 | 1 | 0 | 2 | 5 | 5 | 0 | 1 | 0 | 0 | 2 | 1 | 0 | 4 | 4 |
| - > 50,000 | 0 | 0 | 1 | 4 | 1 | 0 | 1 | 0 | 1 | 1 | 0 | 0 | 0 | 2 | 6 | 0 | 0 | 2 |
| Number of pediatric positive COVID-19 cases seen in ED (total for all centers per country) | 7** | 16 | 0 | 34 | 13** | 2 | 11 | 4 | 48 | 2 | 0 | 1 | 0 | 12 | 26 | 0 | 14** | 2** |
| Time from first COVID case | 3-4 weeks | 3-4 weeks | 3-4 weeks | ≥ 4 weeks | 3-4 weeks | 3-4 weeks | 2-3 weeks | 3-4 weeks | ≥ 4 weeks | 2-3 weeks | 3-4 weeks | 1-2 weeks | 3-4 weeks | 2-3 weeks | ≥ 4 weeks | ≥ 4 weeks | 3-4 weeks | ≥ 4 weeks |
| Number of total COVID-19 confirmed cases | ≥1,000<br><10,000 | ≥100<br><1,000 | ≥100<br><1,000 | ≥10,000 | ≥1,000<br><10,000 | ≥100<br><1,000 | ≥100<br><1,000 | ≥100<br><1,000 | ≥10,000 | ≥10<br><100 | ≥10<br><100 | ≥10<br><100 | ≥1,000<br><10,000 | ≥100<br><1,000 | ≥10,000 | ≥1,000<br><10,000 | ≥1,000<br><10,000 | ≥1,000<br><10,000 |
| N° of total COVID-19 confirmed deaths | ≥10<br><100 | <10 | <10 | ≥100<br><1,000 | ≥10<br><100 | <10 | <10 | <10 | ≥1,000 | <10 | <10 | <10 | ≥100<br><1,000 | <10 | ≥1,000 | ≥10<br><100 | ≥10<br><100 | ≥100<br><1,000 |
| Number of total pediatric COVID-19 confirmed cases | ≥10<br><100 | ≥10<br><100 | ≥10<br><100 | ≥1,000<br><10,000 | ≥100<br><1,000 | ≥10<br><100 | ≥10<br><100 | <10 | ≥100<br><1,000 | <10 | <10 | <10 | <10 | ≥10<br><100 | Not reported | ≥10<br><100 | ≥100<br><1,000 | Not reported |

ED=Emergency Department; PED= Pediatric Emergency Department; UK= United Kingdom
\* referral ED for children

Approximately one third of centers (34%) did not have an ED contingency plan for pandemics **(Table 2)**, irrespective of time interval from first COVID case, number of confirmed cases or ED setting. The majority of centers (76%) had not experienced mass casualty disasters or pandemics during the past 5 years and 36% had never had simulations for such events. Nearly all institutions had established a formal ED management plan for suspected/confirmed pediatric COVID-19 cases by March 20^th^, with daily updates in 69% of centers. Surge capacity for pediatric suspected COVID-19 cases was highly variable between centers at an ED, admission ward and intensive care level and was not proportional to the ED pediatric annual census for any of these settings.

**Table 2.**
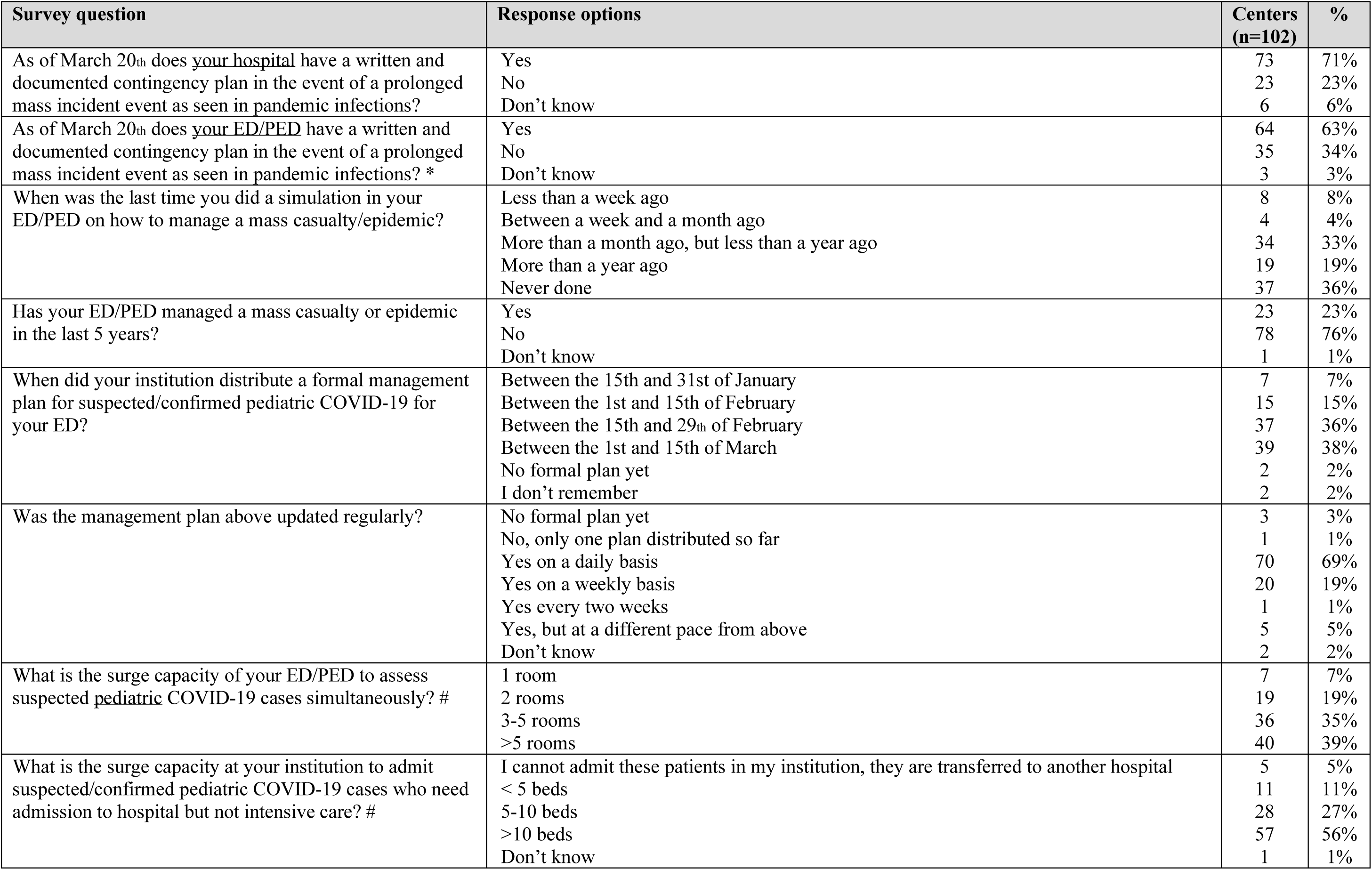
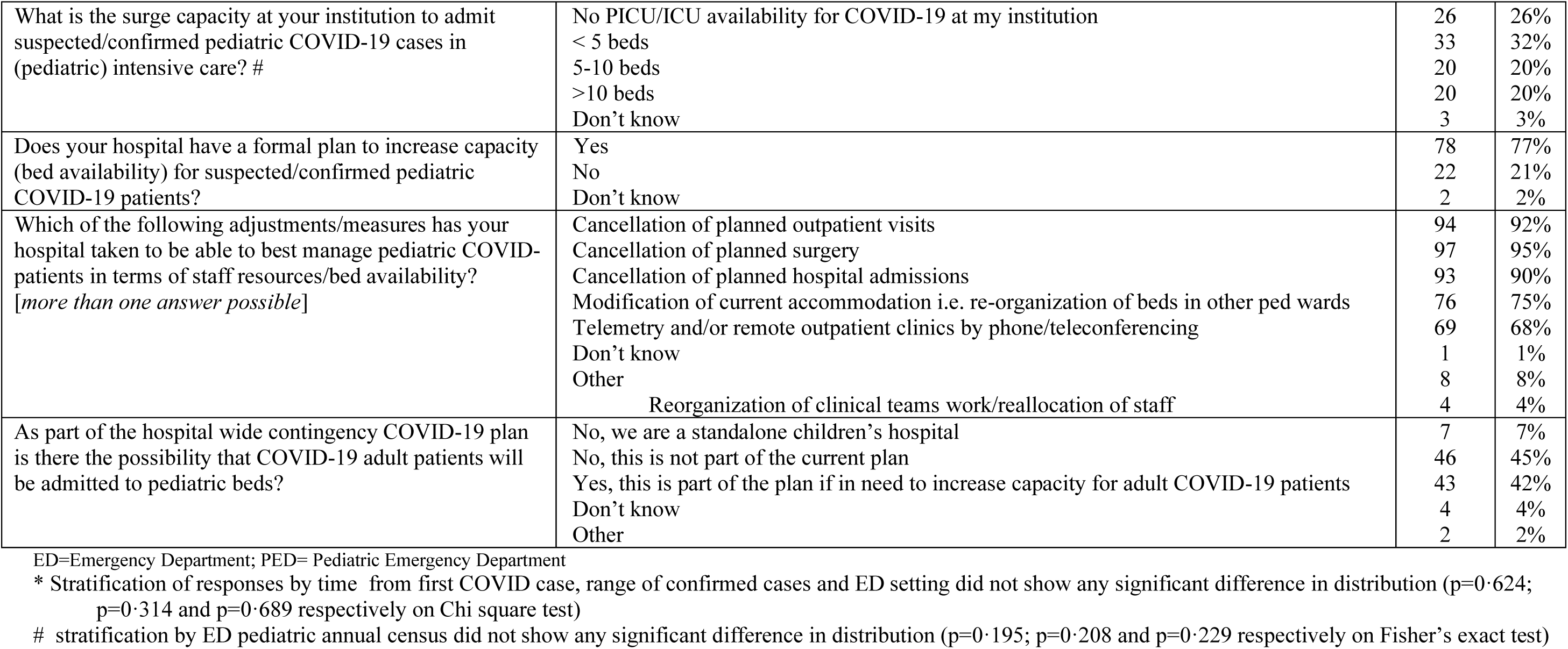
Contingency plans, guidelines and capacity

In one fifth of the institutions there was no intensive care availability for pediatric COVID-19 patients. Plans to increase capacity widely varied between centers. Establishment of a pre-triage and personal protective equipment (PPE) training was also highly variable, as was the use of PPE at pre-triage and for patient assessment. Recommended PPE use for patients was more consistent across centers. Recommended duration of filtering masks use was also variable. A shortage of both basic and erosol generating protective PPE items was experienced by nearly two thirds of centers with masks being the items most frequently missing (**Table 3**).

**Table 3.**
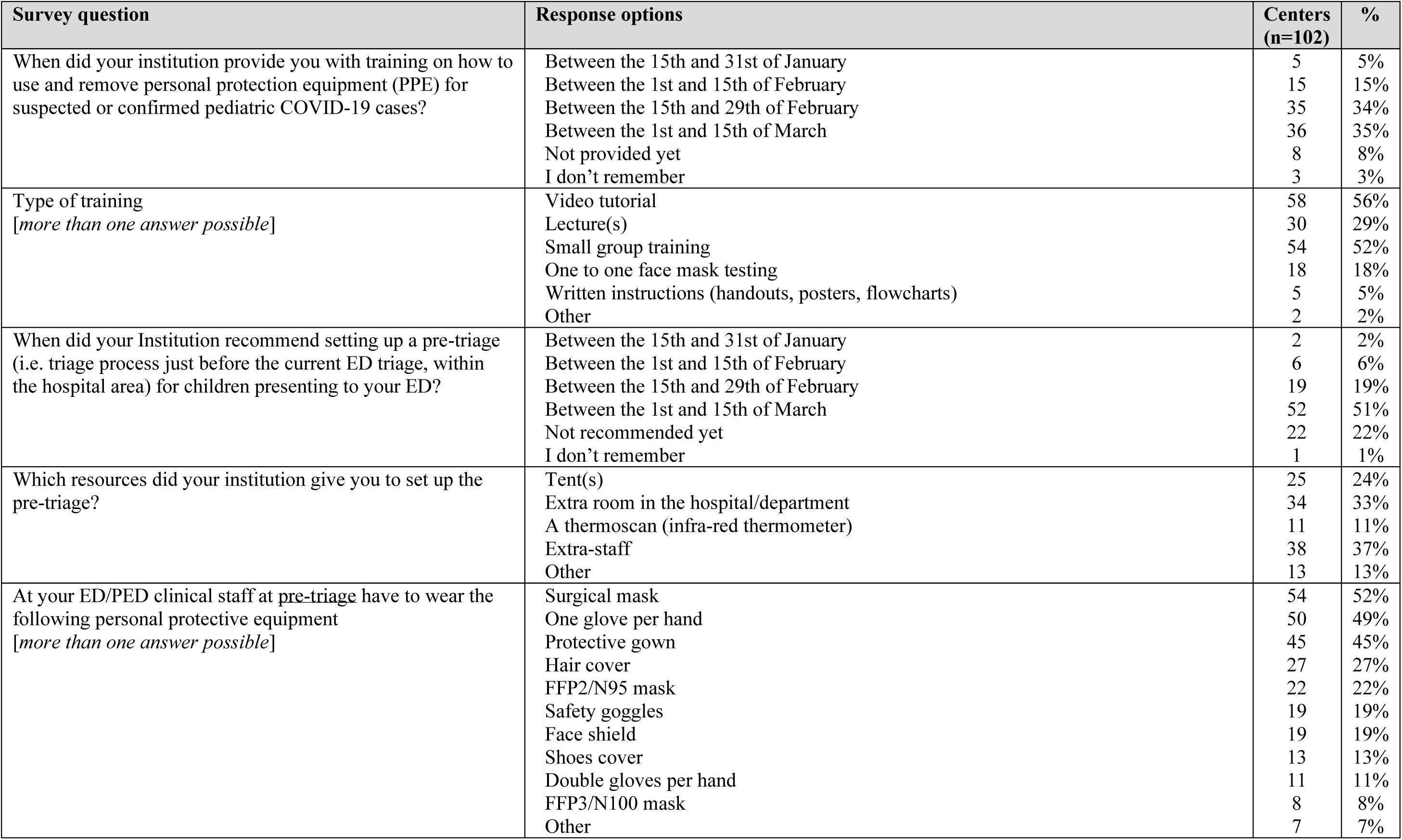
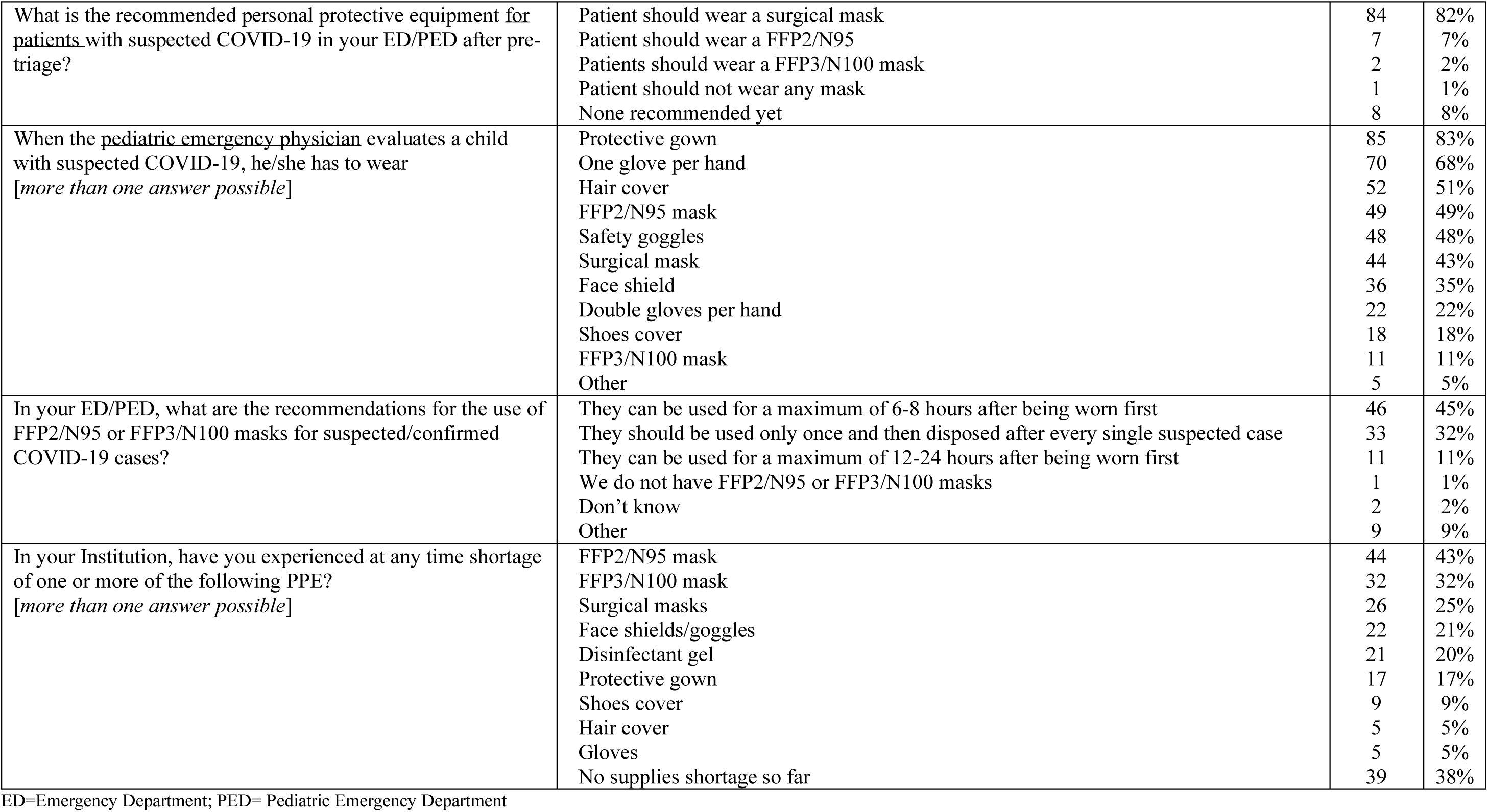
Personal protective equipment and pre-triage

| Survey question | Response options | Centers<br>(n=102) | % |
| --- | --- | --- | --- |
| When did your institution provide you with training on how to use and remove personal protection equipment (PPE) for suspected or confirmed pediatric COVID-19 cases? | Between the 15th and 31st of January<br>Between the 1st and 15th of February<br>Between the 15th and 29th of February<br>Between the 1st and 15th of March<br>Not provided yet<br>I don't remember | 5<br>15<br>35<br>36<br>8<br>3 | 5%<br>15%<br>34%<br>35%<br>8%<br>3% |
| Type of training<br>*[more than one answer possible]* | Video tutorial<br>Lecture(s)<br>Small group training<br>One to one face mask testing<br>Written instructions (handouts, posters, flowcharts)<br>Other | 58<br>30<br>54<br>18<br>5<br>2 | 56%<br>29%<br>52%<br>18%<br>5%<br>2% |
| When did your Institution recommend setting up a pre-triage (i.e. triage process just before the current ED triage, within the hospital area) for children presenting to your ED? | Between the 15th and 31st of January<br>Between the 1st and 15th of February<br>Between the 15th and 29th of February<br>Between the 1st and 15th of March<br>Not recommended yet<br>I don't remember | 2<br>6<br>19<br>52<br>22<br>1 | 2%<br>6%<br>19%<br>51%<br>22%<br>1% |
| Which resources did your institution give you to set up the pre-triage? | Tent(s)<br>Extra room in the hospital/department<br>A thermoscan (infra-red thermometer)<br>Extra-staff<br>Other | 25<br>34<br>11<br>38<br>13 | 24%<br>33%<br>11%<br>37%<br>13% |
| At your ED/PED clinical staff at <u>pre-triage</u> have to wear the following personal protective equipment<br>*[more than one answer possible]* | Surgical mask<br>One glove per hand<br>Protective gown<br>Hair cover<br>FFP2/N95 mask<br>Safety goggles<br>Face shield<br>Shoes cover<br>Double gloves per hand<br>FFP3/N100 mask<br>Other | 54<br>50<br>45<br>27<br>22<br>19<br>19<br>13<br>11<br>8<br>7 | 52%<br>49%<br>45%<br>27%<br>22%<br>19%<br>19%<br>13%<br>11%<br>8%<br>7% |
| What is the recommended personal protective equipment for patients with suspected COVID-19 in your ED/PED after pre-triage? | Patient should wear a surgical mask<br>Patient should wear a FFP2/N95<br>Patients should wear a FFP3/N100 mask<br>Patient should not wear any mask<br>None recommended yet | 84<br>7<br>2<br>1<br>8 | 82%<br>7%<br>2%<br>1%<br>8% |
| When the pediatric emergency physician evaluates a child with suspected COVID-19, he/she has to wear<br>*[more than one answer possible]* | Protective gown<br>One glove per hand<br>Hair cover<br>FFP2/N95 mask<br>Safety goggles<br>Surgical mask<br>Face shield<br>Double gloves per hand<br>Shoes cover<br>FFP3/N100 mask<br>Other | 85<br>70<br>52<br>49<br>48<br>44<br>36<br>22<br>18<br>11<br>5 | 83%<br>68%<br>51%<br>49%<br>48%<br>43%<br>35%<br>22%<br>18%<br>11%<br>5% |
| In your ED/PED, what are the recommendations for the use of FFP2/N95 or FFP3/N100 masks for suspected/confirmed COVID-19 cases? | They can be used for a maximum of 6-8 hours after being worn first<br>They should be used only once and then disposed after every single suspected case<br>They can be used for a maximum of 12-24 hours after being worn first<br>We do not have FFP2/N95 or FFP3/N100 masks<br>Don't know<br>Other | 46<br>33<br>11<br>1<br>2<br>9 | 45%<br>32%<br>11%<br>1%<br>2%<br>9% |
| In your Institution, have you experienced at any time shortage of one or more of the following PPE?<br>*[more than one answer possible]* | FFP2/N95 mask<br>FFP3/N100 mask<br>Surgical masks<br>Face shields/goggles<br>Disinfectant gel<br>Protective gown<br>Shoes cover<br>Hair cover<br>Gloves<br>No supplies shortage so far | 44<br>32<br>26<br>22<br>21<br>17<br>9<br>5<br>5<br>39 | 43%<br>32%<br>25%<br>21%<br>20%<br>17%<br>9%<br>5%<br>5%<br>38% |

ED=Emergency Department; PED= Pediatric Emergency Department

Contagion of healthcare workers was frequently reported at an institution level (69%), but less so at the ED level (25%). Only 18% of sites endorsed a periodic active surveillance of ED staff. Disposition of healthcare workers who had been in close contact with a confirmed COVID-19 case varied between centers, with approximately one third allowing staff to work while asymptomatic and one third recommending quarantine at home. Overall, ED physicians shift work had been re-arranged in nearly two thirds of centers with variable adjustments including both increase and reduction in staff, as well as different shift schemes to prevent cross-infection among staff (**Table 4**).

**Table 4.** ED Staff safety and service sustainability

| Survey question | Response options | Centers<br>(n=102) | % |
| --- | --- | --- | --- |
| Does your institution have an active surveillance plan to test ED staff on a regular basis for COVID-19? | Yes<br>No<br>Don't know | 18<br>82<br>2 | 18%<br>80%<br>2% |
| In your Institution, are you aware of any COVID-19 case in a healthcare worker? | Yes<br>No | 71<br>31 | 69%<br>31% |
| In your Institution, if a healthcare worker has been in contact (without personal protection devices) with a confirmed case of COVID-19 | He/she must be tested and in the meantime be in quarantine<br>He/she must be tested and in the meantime can work with a surgical mask<br>He/she must be placed in quarantine without being tested<br>If he/she has no symptoms, can continue to work without being tested<br>Other<br>Don't know | 31<br>16<br>10<br>36<br>7<br>2 | 31%<br>15%<br>10%<br>35%<br>7%<br>2% |
| Has any of your ED/PED staff tested positive for COVID-19? | Yes<br>No<br>Don't know | 25<br>76<br>1 | 25%<br>74%<br>1% |
| Have staff physicians shifts been rearranged to face the COVID-19 emergency for pediatric cases? | Yes<br>No | 64<br>38 | 63%<br>37% |
| How were shifts re-arranged? (data on n=64 positive responses to previous question) | We increased the number of ED consultants for children<br>We reduced the number of ED consultants for children dividing them in teams to have a spare team in case someone gets infected/needs quarantine<br>Staff physicians are organized in groups which rotate in the same way (to prevent infection among staff)<br>Other<br>Staff re-allocation to other services/roles<br>Spare staff for replacement of colleagues who may get sick | 14<br>16<br>20<br>14<br>3<br>3 | 22%<br>25%<br>31%<br>14% |

ED=Emergency Department; PED= Pediatric Emergency Department

EDs limited caregivers/parents presence to only one person in the majority of centers (84%) and reorganized patient flow to accommodate suspected cases in separate dedicated areas. Fewer than 20% of EDs had isolation rooms with negative pressure. While most EDs performed swab testing for SARS-CoV-2 (78%), there was wide variability on how the test was performed. However, in the majority of centers (75%), asymptomatic children with a history of close contact, who could be otherwise discharged, were not tested in the ED. At most sites suspected cases who were tested, but were fit for discharge, were sent home and swab results communicated to the family when they became available. In cases of positive test results in discharged patients, half of the centers could count on specific outpatient services to provide telephone follow-up. Most EDs experienced a significant reduction in pediatric presentations, by more than 50% in half of the centers (**Table 5**). A longer time since first case was significantly associated with a larger reduction in the number of presentations (p=0.003; **Figure 1)**.

**Table 5.** Logistics and organization of ED management

| Survey question | Response options | Centers (n=102) | % |
| --- | --- | --- | --- |
| In your ED/PED what is the policy for parental/caregiver presence of children with suspected COVID-19? | Both parents/caregivers are allowed to be with the child<br>Only one parent/caregiver is allowed to be with the child<br>There are no rules that establish the number of caregivers allowed in the ED | 13<br>86<br>3 | 13%<br>84%<br>3% |
| What is the patient flow for a suspected COVID-19 in your ED/PED after pre-triage? | Patient is taken directly to an isolation room, with negative pressure<br>Patient is taken directly to an isolation room, but with no negative pressure<br>Patient is taken directly in a usual visit room<br>Patient waits in the usual waiting room<br>Patient waits in a dedicated waiting room for suspected COVID-19<br>Other | 14<br>38<br>7<br>1<br>36<br>6 | 14%<br>37%<br>7%<br>1%<br>35%<br>6% |
| Where does the Pediatric Emergency physician evaluate a child with suspected COVID-19? | in an isolation room, with negative pressure<br>in an isolation room, but with no negative pressure<br>in a usual visit room<br>Other | 18<br>63<br>18<br>3 | 17%<br>62%<br>18%<br>3% |
| If you evaluate a child with suspected COVID-19, do you perform nasal/pharyngeal swab(s) for SARS-CoV-2 in the ED/PED? | Yes<br>No<br>Don't know | 79<br>22<br>1 | 78%<br>21%<br>1% |
| If you decide to perform nasal/pharyngeal swab for SARS-CoV-2, which swabs do you/will you take? | The nasal swab only<br>The pharyngeal swab only<br>Both (nasopharyngeal swab) with one swab stick and one tube (first pharynx first and then nose)<br>Both (nasopharyngeal swab) with two separate swab sticks in two separate tubes<br>Don't know | 23<br>17<br>41<br>20<br>1 | 22%<br>17%<br>40%<br>20%<br>1% |
| If you perform a nasal/pharyngeal swab for SARS-CoV-2 in the ED/PED to a clinically stable child (who would not otherwise require admission) | I must keep the child in a dedicated isolation room until I receive the swab result<br>I can discharge the child home and I communicate the family the result when available<br>I have to admit the child to a regular pediatric ward until I receive the swab result<br>Children who do not need admission are not tested for SARS-CoV-2 in my ED<br>Other | 6<br>78<br>2<br>15<br>1 | 6%<br>75%<br>2%<br>15%<br>1% |
| If your only criterion for suspected COVID-19 is close contact with a confirmed COVID-19 case and the child is otherwise asymptomatic and well, do you test/will you test the child in the ED/PED for SARS-CoV-2? | No, they are discharged home for isolation (quarantine)<br>No, they are redirected to a screening clinic where swabs are done on asymptomatic patients<br>Yes always<br>Yes sometimes, depending on other circumstances | 77<br>7<br>9<br>9 | 75%<br>7%<br>9%<br>9% |
| If you have a confirmed pediatric COVID-19 case | I must admit the child for 14 days independently of his/her clinical conditions<br>I can discharge the child based on his/her clinical conditions, as I do for any patient, recommending home quarantine<br>Other | 5<br>87<br>6 | 5%<br>85%<br>6% |
|  | Don't know | 4 | 4% |
| In your city, are there specific outpatient services that provide phone follow-up of discharged children with confirmed pediatric COVID-19 cases? | Yes | 49 | 48% |
|  | No | 45 | 44% |
|  | Don't know | 8 | 8% |
| Are you experiencing a reduction in number of pediatric visits in your ED/PED since the start of the COVID-19 outbreak in your country? | No | 7 | 7% |
|  | Yes, up to 25% | 14 | 14% |
|  | Yes, up to 50% | 27 | 26% |
|  | Yes, by more than 50% | 52 | 51% |
|  | Don't know | 2 | 2% |

ED=Emergency Department; PED= Pediatric Emergency Department

**Figure 1.**
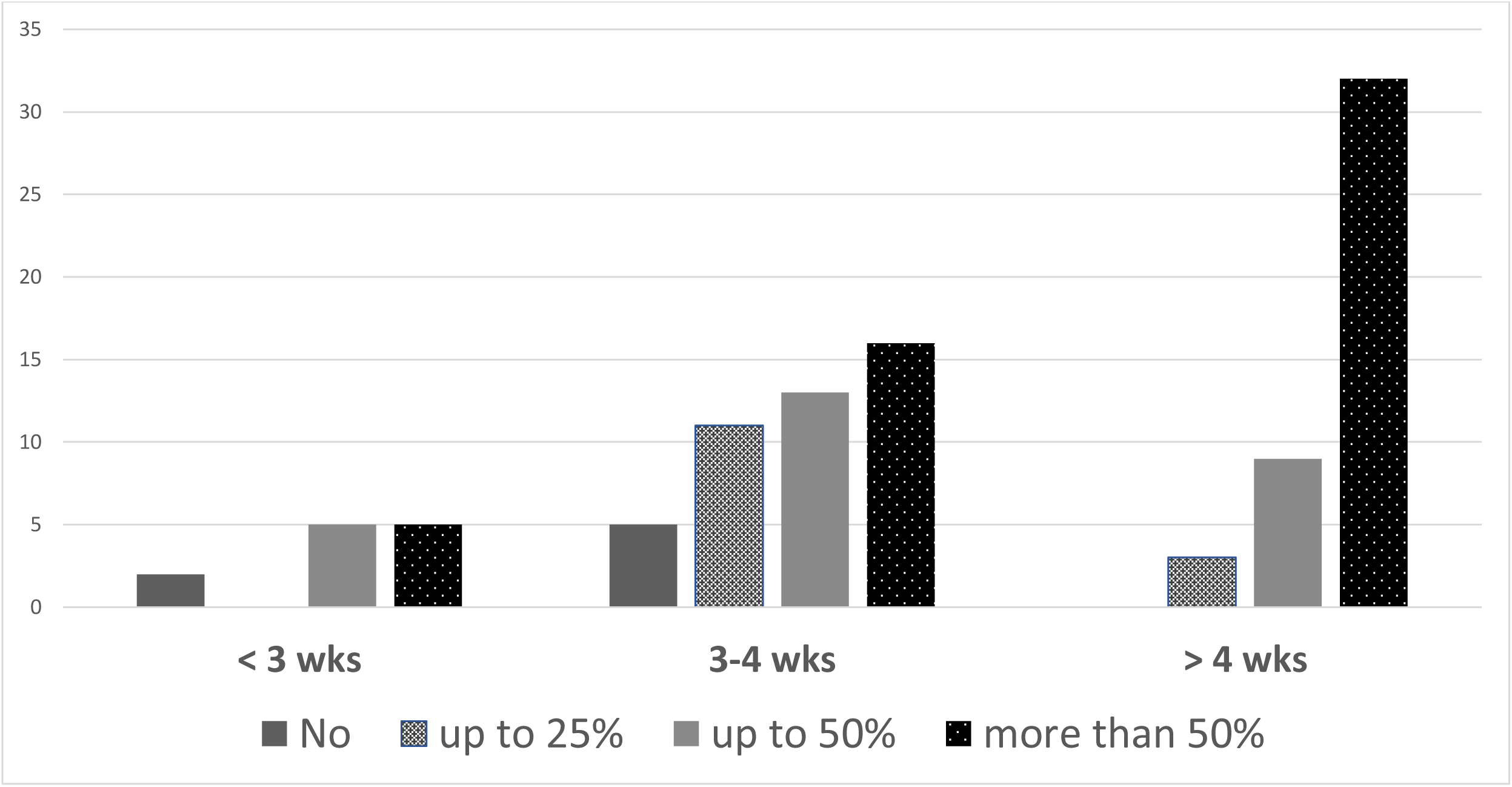
Reported reduction in pediatric ED visits by time since first reported COVID-19 case (based on country of origin)

Local/national health authority documents, hospital policy/infectious disease expertise and websites of international organizations, and published article from China received the higher rating scores as useful sources to inform preparedness and management of pediatric COVID-19 (**Figure 2S**). Overall 46% of centers agreed (36%) or strongly agreed (10%) about the statement “My hospital was ready and prepared to handle COVID-19 at the time the outbreak started in our country” and 54% agreed (39%) or strongly agreed (15%) when the statement was referred to ED pediatric care.

## LIMITATIONS

The results of our study should be interpreted in the light of its limitations. Although we included a large number of European countries, our survey does not provide a pan-European perspective. However, this is the first European dataset that provides a detailed snapshot of pediatric emergency care from within the pandemic, at a more granular level than any institutional channel has been able to provide so far. While the pandemic evolves in each country and accompanying adjustments are made, a repeat focused survey will capture the dynamic progress made from an organizational and operational perspective. We arbitrarily decided, as a research team, the number of centers to be included in each country to ensure a balanced representativeness and to obtain timely completion of the survey. The participating centers represent a subset of EDs caring for children in Europe and include referral centers for children, thus our findings may not be generalisable across different settings. Although some countries exceeded the expected number of recruited centers, we were able to obtain a reasonable balance in terms of country representativeness. In addition, the objective of this survey was to explore common challenges and generalizable learning points and not to compare responses between countries.

## DISCUSSION

Our survey provides a snapshot of preparedness and response of EDs caring for children from 17 European countries and a European associated country at one month after the COVID-19 outbreak started in Northern Italy. Overall, the findings of our study show high variation in time and in level of organisational responses to COVID-19 of EDs caring for children across Europe. While participating countries were at different stages in the outbreak spread the different pace in the pandemic advancement represents an opportunity for healthcare systems to learn from each other. This may ensure a more rapid response in terms of implementation of infection prevention and control measures within healthcare in those countries that lag behind the spread wave. This is important at all levels of care within an integrated health care system, but it is paramount for frontline services such as EDs.^18,19^ Our findings show that a written and documented contingency plan was still missing in approximately one third of centers one month after the onset of the outbreak in Europe. While the majority had not faced an epidemic or a mass casualty event in the past five years, nearly 40% had never run a simulation on how to manage such a crisis in the ED. Although children have shown to be relatively spared from this pandemic,^5,6,20^ timely preparation and appropriate response are essential to minimize the transmission of the infection to both patients and healthcare professionals. Healthcare facilities have played an unwillingly significant role in increasing viral transmission in this pandemic.21 For physicians taking care of children in the ED, COVID-19 has rightfully been defined a logistic rather than a clinical emergency, and the low numbers of patients seen in participating EDs confirms this.^22^ While by March 20^th^ nearly all participating centers had received a formal plan for the management of pediatric suspected/confirmed COVID-19 cases, many faced common challenges: the lack of unequivocal definition of pediatric suspected cases and the need of continuous adjustments secondary to the rapid change of definitions and management plan; the late training in PPE use and shortage in PPE supplies; the need for extra-resources to set up a pre-triage; the re-arrangement of staff shift work to minimize infection spread or to cover for sick colleagues; the lack of negative pressure isolation rooms (if new hospitals are being built, the pandemic perspective isolation rooms should be used); the lack of outpatient services to follow up discharged children with confirmed COVID-19, with possible avoidable representations to the ED; the possibility to admit adult COVID-19 patients into pediatric beds; the difficult balance of resource use.

A striking finding of our point prevalence survey was the wide variation in reported PPE use at pre-triage and for the assessment of suspected COVID-19 cases. While the survey question might have been misinterpreted with respect to assessment by the emergency physician, as to whether or not this included erosol-generating procedures, suboptimal reported practice still emerged from responses. Appropriate PPE use is paramount for staff safety and to reduce the risk of viral transmission.^23,24^ Although tracheal intubation, manual ventilation or non-invasive ventilation are rarely needed for pediatric COVID-19 patients,^5,6,20^ nearly 80% of participating EDs performed swabbing, which is classified as an erosol-generating procedure.

The PPEs recommended by the interim guidance of the European Center for Disease Prevention and Control and the World Health Organization for health care professionals performing erosol-generating procedures are gown, respirator (N95 or FFP2 standard or equivalent), gloves, eye protection (goggles or face shield) and apron, while those providing direct care to COVID-19 patients should wear a gown, surgical mask, gloves and eye protection. Healthcare workers at triage should maintain special distance of at least 1 metre and provide the patient with a medical mask (if tolerated); no PPE is required if preliminary screening does not involve direct patient contact.^25,26^ Nearly half the centers reported a shortage of PPE, most often masks. PPE use should be maximized to avoid shortage of supplies, which ultimately exposes staff and the broader community to an increased transmission risk. One third of respondents stated that respirators (N95/FFP2 or FFP3/N100) are disposed of after the assessment of each suspected case. This practice may contribute to shortage of supply, as the same respirator could be used for more than one patient, as long as it is not damaged or soiled.

A high percentage of centers reported infection in staff members. Unfortunately, infection of healthcare workers has been reported as a major threat to the sustainability of healthcare in this pandemic.^23^ In fact, the disposition of healthcare professionals who had been in close contact with a confirmed COVID-19 case varies between centers, probably because of concerns regarding service provisions.

Implementation of appropriate PPE use can be easily done and should occur in a timely manner. This is in contrast to barriers related to structural limitations and constraints affecting the organization of ED patient flow and isolation capacity, which may be difficult to overcome in a short time frame. Infection control measures were more consistently reported in the survey, including re-arrangement of ED patient flow, changing of staff work shift to optimize resource utilization, reduction in the number of care givers allowed with the child and home quarantine for confirmed COVID-19 pediatric cases fit for discharge.

Another interesting finding from our survey is the substantial reduction in pediatric ED presentations during the pandemic, which greatly helped centers with more limited isolation capacity better manage suspected COVID-19 cases. Centers from countries with a longer time since first case experienced higher reductions in the number of ED presentations. Parents’ fear of contagion in a healthcare environment, improved hygiene measures, reduced community transmission of communicable diseases, reduced opportunities to sustain injuries owing to the strict containment measures enforced by governments, and reduction in stress-related functional diseases may be the reasons underlying this phenomenon. Reports from previous epidemics also showed an overall decrease in PED attendances.^27,28^ The MERS outbreak had resulted in a significantly higher proportion of high acuity ED pediatric presentations and an increase in delayed presentations.^9,29^ High levels of care for children presenting with non-COVID-19 related acute illnesses, including children with complex medical needs, should be maintained during the pandemic.

Despite its limitations, the provision of a timely report on preparedness and response in pediatric emergency care during the pandemic is useful to inform practice and policymakers to properly re-organize health systems while the crisis is still evolving. It provides an accurate objective historical dataset from which lessons can be learnt for the future, including for adult practice. The collaboration of the REPEM and PERUKI European networks was instrumental to ensure wide representation of European countries and timely completion of this multinational point prevalence survey. The data provided highlights the importance of European multinational research collaborations to provide the best care to children in the frontline.

In summary, we identified variability and gaps in preparedness and response to the COVID-19 epidemic across European referral EDs for children at one month since the start of the outbreak in Northern Italy. Early availability of a written and documented contingency plan including detailed infection control measures, the provision of simulation training, appropriate use of PPE, and appropriate isolation facilities emerged as key factors that should be optimized to improve the preparedness and inform responses to future pandemics.

## Data Availability

The data that support the findings of this study are available from the corresponding author, [SB], upon reasonable request.

## Author contributions

Silvia Bressan: Conceived the study, designed the study, obtained, analysed and interpreted the data, wrote the initial draft of the paper, gave final approval to be published, and agreed to be accountable for all aspects of the work.

Santiago Mintegi: Conceived the study, designed the study, interpreted the data, critically revised the draft of the paper, gave final approval to be published, and agreed to be accountable for all aspects of the work.

Danilo Buonsenso designed the draft of the survey.

Danilo Buonsenso, Niccolo’ Parri, Ruth Farrugia, Ruud Nijman, Rianne Oostenbink, Luigi Titomanlio, Ian Maconochie, Damian Roland and Liviana Da Dalt: Designed the study, contributed to the interpretation of the data, drafted or revised it critically, gave final approval to be published, and agreed to be accountable for all aspects of the work.

## Contributors’ list

*Country leads list (country leads collaborated closely with the research team and were instrumental for the dissemination and completion of the survey)*. Said Hachimi Idrissi, UZ Gent, Brussel, Belgium; Marianne Sjølin Frederiksen, Copenhagen University Hospital, Herlev, Denmark; Ulle Uustalu, Tallinn Children's Hospital, Estonia; Cheron Gerard, Necker enfants malades, France; Florian Hoffmann, Dr. von Hauner University Children's Hospital Munich, Germany; Valtyr Thors, Children’s hospital Iceland. Landspitali University Hospital, Iceland; Michel J Barrett, Children's Health Ireland at Crumlin, Ireland; Itai Shavit, Rambam Health Care Campus, Isrel; Zanda Pucuka, BKUS Children clinical university hospital, Latvia; Lina Jankauskaite, Hospital of Lithuanian University of Health Sciences Kauno Klinikos, Lithuania; Correct spelling:Patrícia Mação, Hospital Pediátrico, Centro Hospitalar e Universitário de Coimbra, Portugal; Ioannis Orfanos, Skane University Hospital, Sweden; Laurence Lacroix, Geneva University Hospitals, Switzerland

## Acknowledgments

The authors would like to acknowledge all the respondents to the survey and who gave permission for their names to be included in the acknowledgment section of this paper.

The full list of acknowledgments is available upon request.

## Notes

### Competing Interest Statement

The authors have declared no competing interest.

### Funding Statement

No external funding was received for this work

